# High seroprevalence for SARS-CoV-2 among household members of essential workers detected using a dried blood spot assay

**DOI:** 10.1101/2020.06.01.20119602

**Authors:** Thomas W. McDade, Elizabeth M. McNally, Aaron S. Zelikovich, Richard D’Aquila, Brian Mustanski, Aaron Miller, Lauren A. Vaught, Nina L. Reiser, Elena Bogdanovic, Katherine S. Fallon, Alexis R. Demonbreun

**Affiliations:** Department of Anthropology and Institute for Policy Research, Northwestern University; Canadian Institute for Advanced Research, Child and Brain Development Program; Center for Genetic Medicine, Northwestern University; Division of Cardiology, Department of Medicine, Northwestern University Feinberg School of Medicine; Department of Biochemistry and Molecular Genetics; Division of Infectious Diseases, Dept. of Medicine, Northwestern University Feinberg School of Medicine; Institute for Sexual and Gender Minority Health and Wellbeing and Department of Medical Social Sciences, Northwestern University.; Department of Pharmacology, Northwestern University Feinberg School of Medicine

**Keywords:** COVID-19, SARS-CoV-2, serological testing, IgG, ELISA, capillary blood, dried blood spots

## Abstract

**Objective:** Serological testing is needed to investigate the extent of transmission of SARS-CoV-2 from front-line essential workers to their household members. However, the requirement for serum/plasma limits serological testing to clinical settings where it is feasible to collect and process venous blood. To address this problem we developed a serological test for SARS-CoV-2 IgG antibodies that requires only a single drop of finger stick capillary whole blood, collected in the home and dried on filter paper (dried blood spot, DBS).

**Methods:** An ELISA to the receptor binding domain of the SARS-CoV-2 spike protein was optimized to quantify IgG antibodies in DBS. Samples were self-collected from a community sample of 232 participants enriched with health care workers, including 30 known COVID-19 cases and their household members.

**Results:** Among 30 individuals sharing a household with a virus-confirmed case of COVID-19, 80% were seropositive. Of 202 community individuals without prior confirmed acute COVID-19 diagnoses, 36% were seropositive. Of documented convalescent COVID-19 cases from the community, 29 of 30 (97%) were seropositive for IgG antibodies to the receptor binding domain.

**Conclusion:** DBS ELISA provides a minimally-invasive alternative to venous blood collection. Early analysis suggests a high rate of transmission among household members. High rates of seroconversion were also noted following recovery from infection. Serological testing for SARSCoV-2 IgG antibodies in DBS samples can facilitate seroprevalence assessment in community settings to address epidemiological questions, monitor duration of antibody responses, and assess if antibodies against the spike protein correlate with protection from reinfection.

## Introduction

Serological testing for SARS-CoV-2 IgG antibodies identifies prior viral exposure and, potentially, immunity. Recent surveys suggest a range of seroprevalence rates, with relatively low rates in much of the US.^1,2^ Optimal surveillance of seroprevalence ideally avoids contact between community members and health care providers and surveyors, since such contact may carry risk of exposure and discourage survey participation. An alternative to venipuncture blood collection is finger stick dried blood spot (DBS) sampling.^3,4^ DBS relies on a finger prick with blood drops captured on filter paper, and DBS can be performed in the home with return of sample by mail. DBS sampling has served as the foundation for nationwide newborn screening programs since the 1960s and is increasingly applied as a minimally-invasive alternative for community health research. The Centers for Disease Control and US Postal Service consider DBS specimens nonregulated, exempt materials for return of samples to laboratories.^5^

A robust and quantitative ELISA was granted Emergency Use Authorization from the FDA; this ELISA measures SARS-CoV-2 antibodies in serum.^6^ This same ELISA was recently used to detect a higher than expected seroprevalence for antibodies to SARS-CoV-2 among health care workers and patients in a pediatric dialysis unit.^7^ We adapted this ELISA to measure IgG antibodies to the receptor binding domain (RBD) of the SARS-CoV-2 spike protein in DBS samples. The RBD is often the target of neutralizing antibodies, although data on the frequency of RBD-binding antibodies with neutralizing activity is still limited.^8^

## Methods

A detailed protocol is supplied in the Supplemental Materials. All research activities were implemented under protocols approved by the institutional review board at Northwestern University (#STU00212457 and #STU00212472).

Participants who previously tested positive for SARS-CoV-2 virus were recruited from the community through direct contact. The remainder of participants were recruited through direct contact and willingness of household member participation. DBS sample collection occurred between April 18 and May 20, 2020. DBS samples from 23 individuals presumed to be negative based on the pre-pandemic date of sample collection (2018) were used as negative controls. Samples were analyzed in duplicate, with the average result reported.

### Statistical Analysis

Statistical analyses were performed with Prism (Graphpad, La Jolla, CA). An unpaired two-tailed t-test was used to compare negative and confirmed positive samples. Error bars represent ± standard error of the mean (SEM). Passing-Bablok regression and bivariate correlation were used to evaluate patterns of statistical association across matched DBS and serum samples.

## Results

CR3022, an antibody with defined affinity to the RBD, was used to validate DBS samples in an ELISA. We observed a strong dose-response between CR3022 concentration and optical density (OD) in wells coated with 2.0µg/mL RBD (**Figure 1A**), with no response in the absence of RBD antigen. Responses were approximately linear up to 6.25µg/mL, with a flattening of response at higher concentrations of CR3022. A CR3022 dilution series generated a calibration curve and determined the lower limit of detection (LLD) as 0.032µg/mL. Intra-assay variability (percent coefficient of variance (%CV)) at the seropositivity threshold of 0.6 μg/mL was 4.23, with clear distinction between positive and negative samples (**Figure 1B**). With one exception, all positive samples were more than 3SD from the mean OD for the negative samples. Intraassay variability was 6.56 and 6.20 %CV at 1.56 and 6.25μg/mL, respectively. Matched DBS and serum samples showed tight agreement (**Figure 1C**).

**Figure 1.**
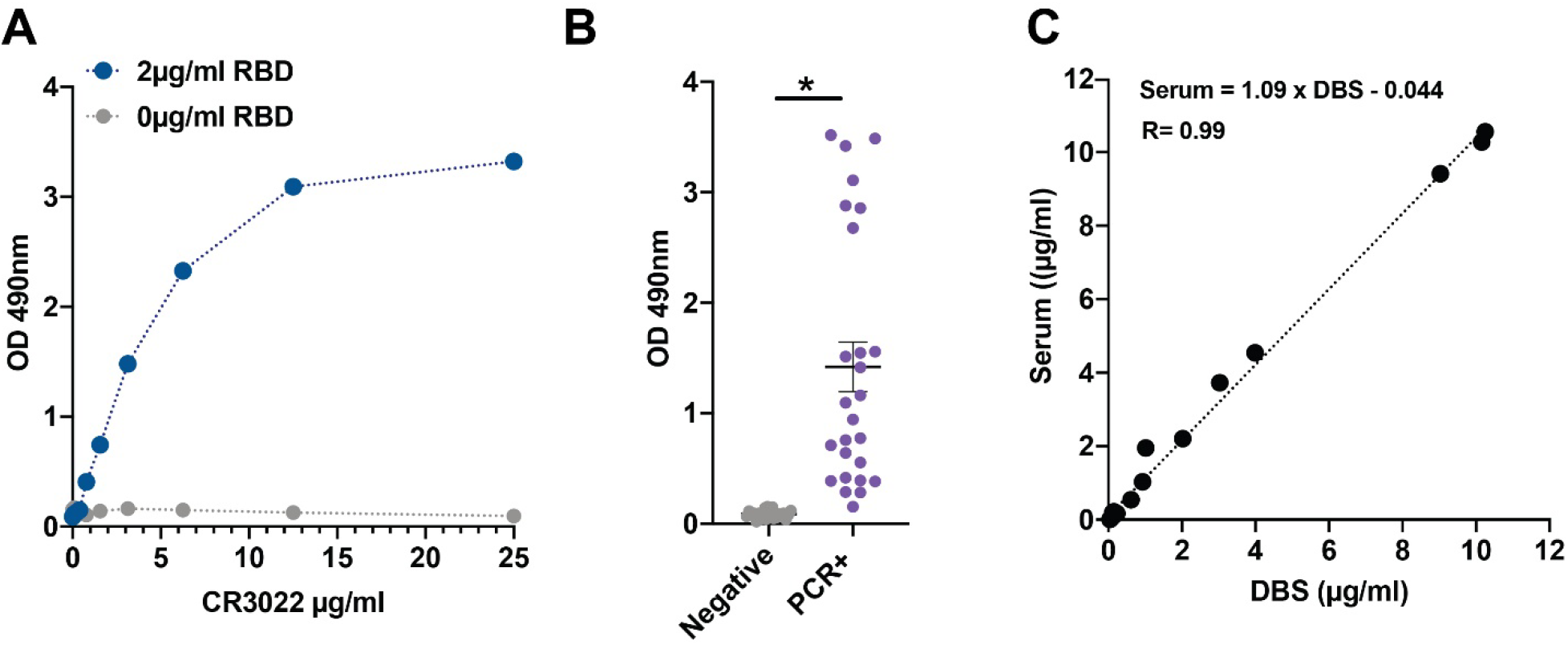
Dried Blood Spot (DBS) ELISA for the receptor binding protein domain of the SARS-CoV-2 spike protein. **A)** The CR3022 antibody has known reactivity to the receptor binding domain protein. Measurements from DBS samples to which known concentrations of CR3022 antibody were added. **B)** Increased signal readily detected in DBS from virus-positive cases. **C)** Near perfect correlation between DBS and serum IgG levels. *p<0.001.

We evaluated IgG levels to RBD protein in 232 DBS samples acquired from the community (**Table 1**). Thirty cases had previous positive viral PCR testing for COVID-19, with 60–85% reporting chills, fatigue, cough, or headache as the most common symptoms. Twenty-seven of 30 were seropositive with IgG concentrations from 0.6μg/ml to 10μg/ml (**Figure 2**). The median time between serological testing and viral PCR testing was 28 days (range 16–43 days). Two participants had low seropositivity (IgG< 0.6μg/ml but > 0.39μg/ml) and both had asymptomatic infections. The single seronegative participant was pregnant, and 52 days after the first viral PCR test she tested positive for virus again.

**Table 1.** Characteristics of study participants in a community-based sample with (PCR+) and without (PCR–) a confirmed case of COVID-19.

|  | PCR+ | PCR- |
| --- | --- | --- |
| Number participants | 30 | 202 |
| Mean age (yrs) | 36 | 37 |
| Age range (yrs) | 24-74 | 18-70 |
| Female | 26 | 101 |
| Male | 4 | 101 |
| Viral PCR test positive | 30 | 0 |
| Viral PCR test negative | 0 | 15 |
| No viral PCR testing | 0 | 187 |
| Mean #days post PCR | 28 | NA |
| Range #days post PCR | 16-43 | NA |

**Figure 2.**
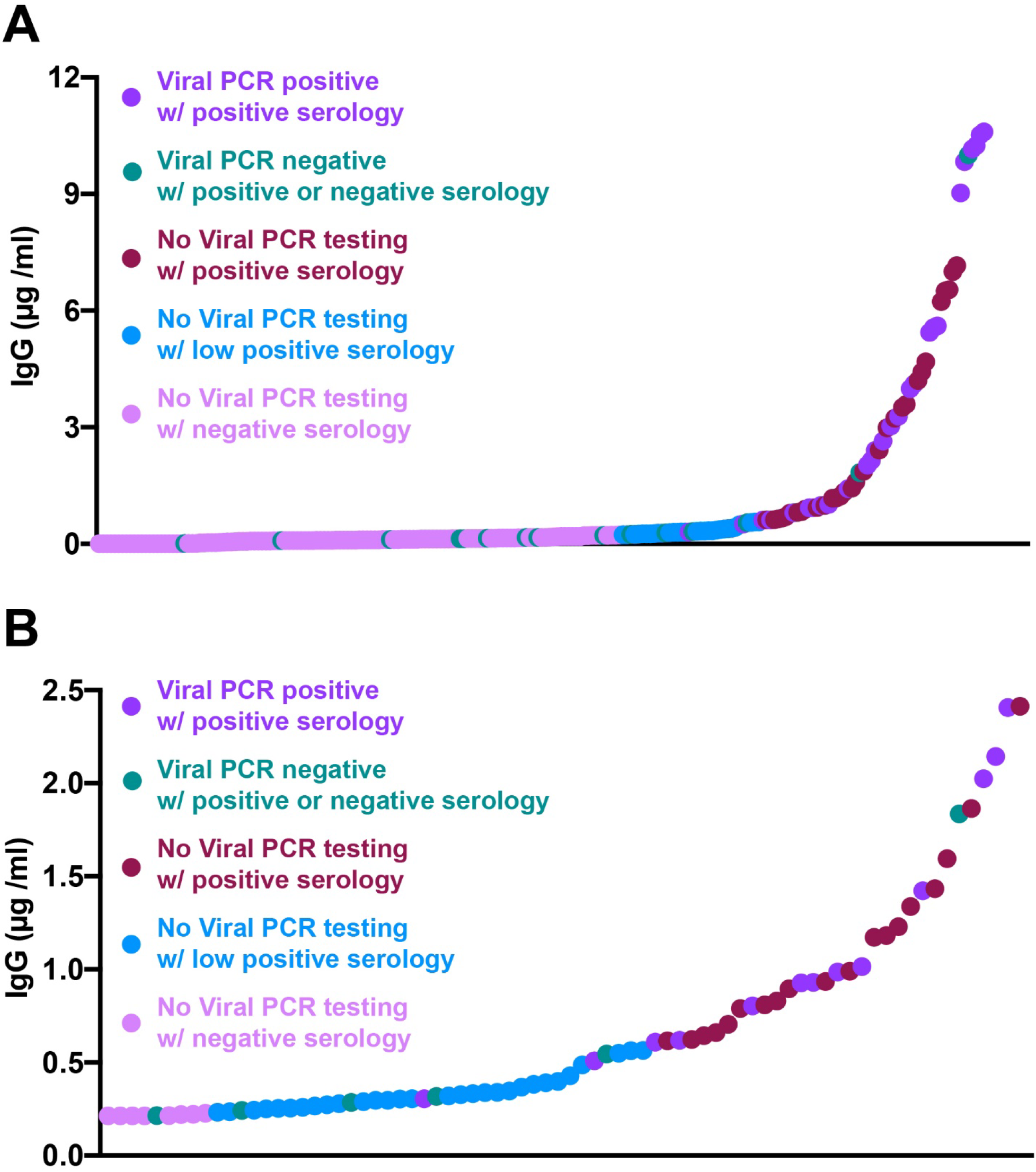
Results from community acquired DBS from April – May 2020. **A)** The range of IgG seropositivity detected in DBS samples collected from 30 known virally infected cases (median 28 days after viral test, range 16–43 days) and 202 without documented COVID-19 infection. **B)** Depicts the lower range of DBS detected seropositivity with those OD greater than 0.6μg/ml considered positive, and less than 0.39μg/ml considered negative. The range in between was considered low seropositive.

Among the remaining 202 participants, none of whom tested positive for SARS-CoV-2 with PCR, 60% reported some COVID-19-like symptoms with headache, fatigue, or rhinorrhea being most common. Fifteen of these participants were PCR tested and determined viral negative; of these, 9 were determined to be seronegative, 4 were low seropositive, and 2 were seropositive in DBS. Only one participant required hospitalization for dyspnea (0.8%), and this individual tested viral negative and was low seropositive.

Follow-up DBS samples were acquired from 17 seronegative and 8 low seropositive participants, 14 days after the original DBS sampling date. IgG to RBD increased in 7 (41%) of the seronegative participants, shifting 3 to seropositive and 4 to low seropositive (**Figure 3**). Six of 8 (75%) low seropositive samples increased IgG to become seropositive.

**Figure 3.**
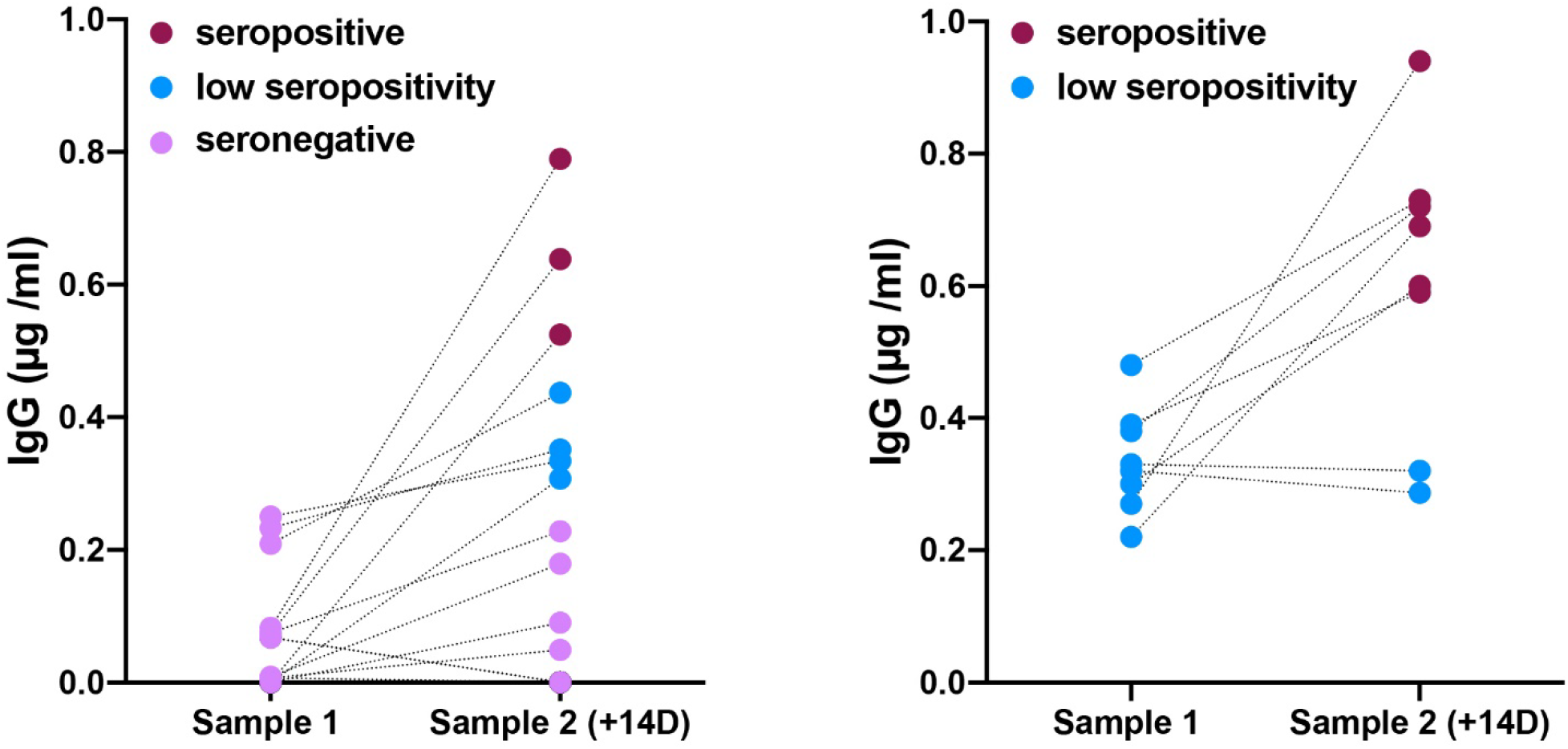
Seroconversion documented in repeat DBS sampling in non-confirmed SARS-CoV-2 subjects. Increased IgG concentrations in 7/17 (41%) seronegative and 6/8 (75%) low seropositive subjects after 14 days.

For the entire 202, IgG concentrations ranged from 0–10μg/ml, with 33 (16%) seropositive, 40 (20%) low seropositive, and 129 (64%) seronegative (**Figure 4A**). We evaluated 30 participants who shared households with individuals confirmed to have SARS-CoV-2 through a viral PCR test. The virus positive index cases in these households were disproportionately healthcare workers or first responders (19/20). Among the 30 household members, 21 (70%) were seropositive, 3 (10%) were low seropositive, and 6 (20%) were seronegative **(Figure 4B)**. A number of participants were from a single location in which a number of healthcare workers tested positive for virus while others were found to be viral negative between March 24 to April 3, 2020. Those healthcare workers that remained asymptomatic were not tested for active virus. We evaluated the seroconversion of 48 household members from 33 essential healthcare worker households, 40–50 days post the initial exposure date **(Figure 4C)**. Participants sharing a household with a known COVID-19 positive healthcare worker had a high seroconversion rate with 9/17 (53%) seropositive, 3/17 (18%) low seropositive, and 5/17 (29%) seronegative. This was in contrast to 10/10 (100%) of household members remaining seronegative when sharing a household with a virus-negative health care worker. Household members that resided with healthcare workers that were not tested for virus were 2/21 (10%) seropositive, 3/21 (14%) low seropositive, and 16/21 (76%) seronegative. These data show a high transmission rate among households of front-line essential workers and the dynamic nature of seroconversion in the community.

**Figure 4.**
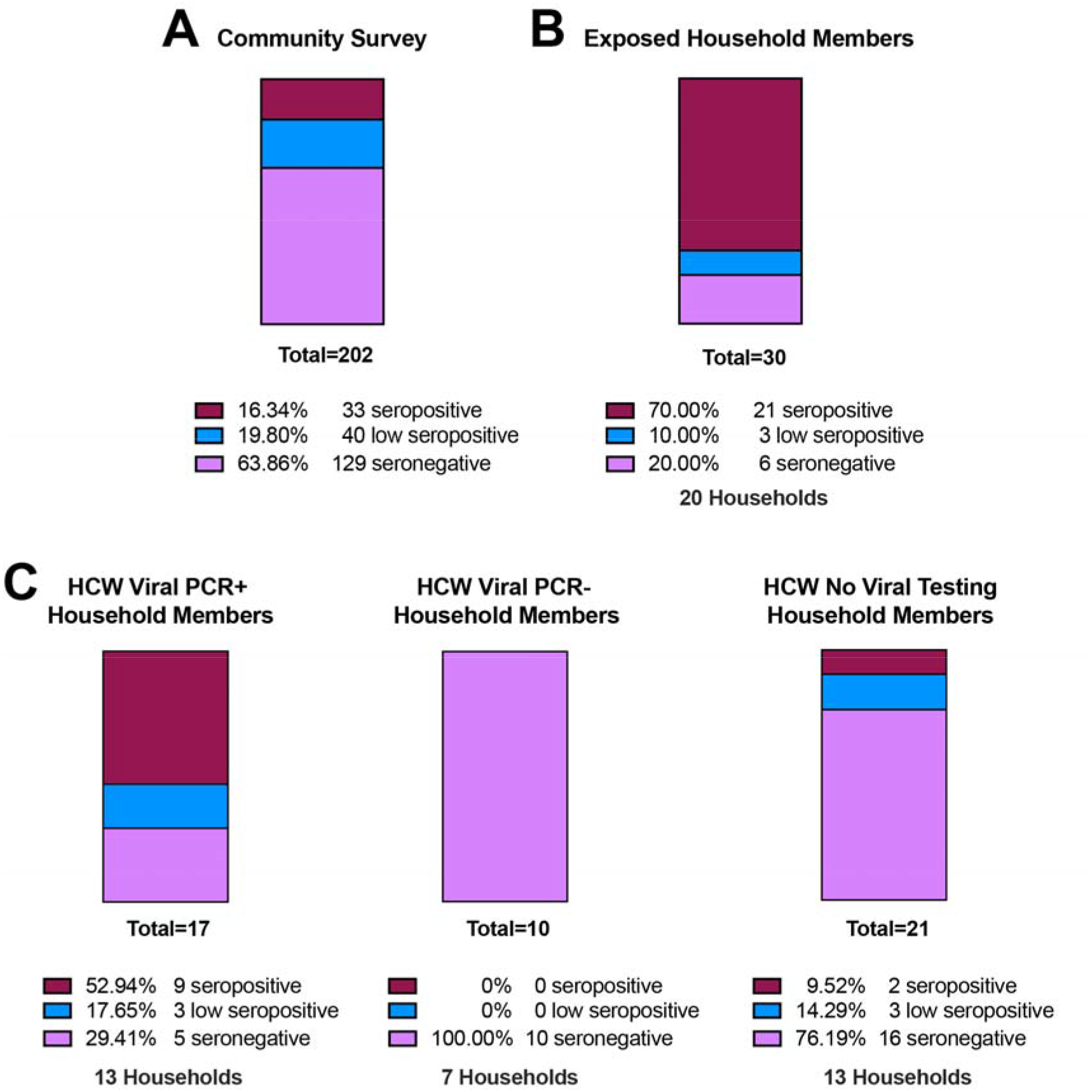
High seroconversion rates in household members of index COVID-19 cases. **A)** Seroprevalence in 202 samples collected from the community, which includes health care workers and first responders, none of which were confirmed SARS-CoV-2 viral positive. **B)** Of 30 COVID-19 exposed household members, 70% were seropositive and 10% low seropositive. **C)** Increased seroconversion in household members of known viral PCR positive healthcare workers (53% seropositive and 18% low positive) compared to 100% seronegative in known viral PCR negative healthcare worker households. Household members of healthcare workers with no viral testing had exposure rates more similar to the community acquired rates.

## Discussion

COVID-19 is now widely acknowledged to have high rates of community spread. Strategic testing in community-based settings is critical for tracking spread of SARS-CoV-2 and for identifying the factors that mitigate transmission. These results demonstrate the feasibility of using DBS samples for serological testing of SARS-CoV-2 IgG antibodies. Results from community sampling support a high rate of household transmission. In addition, based on Hains et al.^7^ and the data here, indeterminate levels of IgG merit retesting and current estimates of seroprevalence may underestimate community spread.

Because of their potential risks to patients, health care workers and first responders generally had priority for viral PCR testing to ascertain acute infection. However, limited supplies of viral testing materials often restricted access to viral PCR testing during March and April 2020, which likely resulted in under-testing and under-reporting of COVID-19 cases. Serological testing is positioned better to inform viral exposure especially during the interval when viral PCR testing was less available or weeks after symptom onset. Furthermore, improved and reliable serological surveys, combined with close clinical follow up, can elucidate whether serological status predicts immune protection from repeat infection or disease. DBS sampling can aid in these surveys.

Advantages of DBS include a low cost and non-invasive approach to blood collection. DBS samples do not require special handling prior to transport and analysis.^3,9^ Antibodies remain stable in DBS for weeks at room temperature. While point-of-care lateral flow immunoassay tests share some of the advantages of DBS sampling, they are difficult to implement in the home and they do not provide the same level of accuracy and quantification as lab-based tests for SARS-CoV-2 antibodies.^10^ Widespread serological testing is needed to more accurately assess viral spread. The observation of high household seroconversion, using an assay restricted to a small component of the viral spike protein, suggests there may be higher exposure to COVID-19 in some locations than previously appreciated.

## Data Availability

Data available upon written request to the corresponding author.

## Acknowledgments

This research was supported with funding from the Canadian Institute for Advanced Research and Center for Genetic Medicine, Northwestern University.

## SUPPLEMENTAL INFORMATION

### Detailed Methods

DBS samples are eluted overnight and transferred to microtiter wells, which were previously coated with SARS-CoV-2 RBD antigen. Goat anti-human IgG-HRP binds to the RBD-antibody complexes in each well, and color forms with the addition of chromogenic substrate. The absorbance of the solution is read at 490nm.

### Reagents and supplies

#### Filter papers

Whatman #903 (Cytiva #10534612). These cards are approved by the FDA as Class II medical devices for diagnostic applications, and the CDC implements an independent quality control analysis of each card lot to confirm thickness, flow-rate, absorbency, and purity.

#### Hole punch

A semi-automatic pneumatic system with 5mm punch (Analytical Sales and Services #327500, Flanders, NJ) was used. Manual punches are also readily available that produce 3/16 inch discs (4.8mm) (e.g., McGill #MCG53600C). A 5 mm punch will contain approximately 5uL of serum.^11,12^

#### Microtiter plates

DBS samples are eluted in Corning 96 Well TC-Treated Microplates (Sigma #CLS3599). For coating with antigen: Immulon 4 HBX polystyrene 96-well flat bottomed, high-binding plates (ThermoFisher Scientific #3855).

#### Coating antigen

The plasmid for the receptor binding domain (RBD) of the spike (S) glycoprotein gene from SARS-CoV-2, Wuhan-Hu-1 (GenBank: MN908947) was generously provided by Dr. Florian Krammer (Mt. Sinai Medical School, NY) and is described previously.^6^ The construct was made by fusing the N-terminal S protein signal sequence to the spike RBD (amino acids 419 to 541) with a C-terminal hexa-histidine tag. The sequence was codon optimized and subcloned into the pCAGGS mammalian expression vector. Recombinant RBD protein was generated by Evotec using standard methods (Princeton, New Jersey). Briefly, EXPI293 cells were transfected with purified, endotoxin free plasmid DNA. Four L of media was purified using IMAC chromatography 72 hours post transfection. The final recovery of purified recombinant RBD protein was ∼195mg in PBS with an endotoxin level at ∼ 1EU/mg, with a purity > 95%.

#### Detection antibody

Anti-Human IgG (Fab specific)-Peroxidase antibody produced in goat (Sigma-Aldrich #A0293).

#### Positive control

IgG antibody to SARS-CoV S protein (CR3022 antibody, Creative Biolabs #MRO-1214LC).

#### Coating buffer

Phosphate buffered saline (1x PBS; Fisher BP24384 or equivalent)

#### Wash buffer

PBS, 0.1% Tween 20 (PBS-T; Fisher BP337-500).

#### Blocking solution

0.1% PBS-T, 3% milk (w/v; American Bio AB10109-01000).

#### Chromogenic substrate

SIGMA*FAST™* OPD (Sigma-Aldrich #P9187). One silver and one gold tablet per 2 plates in 20 mL dH2O.

#### Stop solution

3.0M Hydrochloric acid (HCl).

#### Antibody diluent

0.1% PBS-T, 1% milk (w/v).

### Preparation of materials

DBS calibrators are prepared as follows: 1) Dilute CR3022 antibody to desired concentrations in PBS; 2) Collect EDTA whole blood from a negative donor; 3) Add CR3022 dilutions to aliquots of whole blood, mixing gently to avoid hemolysis; and 4) Transfer with a pipet (65 μL/drop) to labeled #903 cards. Dry overnight at room temperature and store at −23°C. It is important to minimize the volume of CR3022 dilution that is added to whole blood, as large volumes (>5% of total volume) will lower hematocrit and adversely affect how the sample is absorbed on the filter paper.

### Protocol

The 96-well plate is coated with antigen as follows: Dilute RBD to 2 μg/mL in PBS, add 100 μL to each well, seal, and incubate overnight at 4°C. DBS samples are eluted overnight. Punch out one 5.0 mm disc of each DBS calibrator, control, and sample, and place in elution plate. Add 250 μL PBS, seal the plate, and elute overnight at 4°C.

The following day, remove plate and wash 4x with 300 μL PBS-T (BioTek ELx50 or equivalent). Add 200 μL per well 0.1% PBS-T, 3% milk. Block covered at room temperature for two hours. Aspirate.

At one hour, remove DBS samples from refrigerator and rotate at 300 rpm (Heidolph Titramax 101 or equivalent). After removing blocking solution, transfer 100 μL eluate from DBS calibrators, controls, and samples to the coated 96-well plate. Cover the plate and incubate at room temperature for two hours. Wash the plate four times with PBS-T.

Prepare a dilution (1:3000) of detection antibody in 0.1% PBS-T, 1% milk. Add 100 μL working antibody solution to each well. Cover and incubate at room temperature for 60 minutes. Wash four times as before.

Prepare chromogenic substrate by dissolving one set of tablets in 20mL dH2O. Do not add silver tablet until ready to use. Add 100 μL chromogenic substrate to each well. Cover the plate, protect from the light, and incubate for 10 minutes at room temperature. Add 50 μL 3M HCl stop solution to each well. Incubate 5 minutes at room temperature. Read the absorbance (optical density, OD) at 490 nm (BioTek ELx808 or equivalent). OD > 0.6 μg/ml CR3022 is considered positive, less than 0.39μg/ml CR3022 is considered negative, and values between are considered low seropositive. Sample IgG concentration (μg/ml) is calculated from the linear regression of the CR3022 calibration curve.

## Notes

### Competing Interest Statement

The authors have declared no competing interest.

### Author Declarations

Northwestern University Institutional Review Board approved protocols #STU00212457 and #STU00212472.

